# Investigating shared genetic architecture between obesity and multiple sclerosis

**DOI:** 10.1101/2022.12.07.22283195

**Authors:** Ruijie Zeng, Rui Jiang, Wentao Huang, Jiaxuan Wang, Lijun Zhang, Yuying Ma, Yanjun Wu, Meijun Meng, Felix W Leung, Qizhou Lian, Weihong Sha, Hao Chen

**Author notes:** Correspondence: Prof. Felix W Leung, David Geffen School of Medicine, University of California Los Angeles, Los Angeles 90024, California, USA; Prof. Qizhou Lian, Department of Medicine, Queen Mary Hospital, Hong Kong SAR 999077, China; Prof. Weihong Sha, Department of Gastroenterology, Guangdong Provincial People’s Hospital, Guangdong Academy of Medical Sciences, Guangzhou 510080, China; Prof. Hao Chen, Department of Gastroenterology, Guangdong Provincial People’s Hospital, Guangdong Academy of Medical Sciences, Guangzhou 510080, China. These authors contributed equally to this work (Ruijie Zeng, Rui Jiang, Wentao Huang, Jiaxuan Wang).

## Abstract

**Background and aims:** Observational studies have suggested a complex relationship between obesity and multiple sclerosis (MS). However, the role of genetic factors in the comorbidity and whether obesity exist consistent shared genetic relationships with MS, remains unclear. Our study aims to investigate the extent of shared genetic architecture underlying obesity and MS.

**Methods:** Based on genome-wide association studies (GWAS) summary statistics, we investigate the genetic correlation by the linkage disequilibrium score regression (LDSC) and genetic covariance analyzer (GNOVA). The casualty was identified by using bidirectional Mendelian randomization. Linkage disequilibrium score regression in specifically expressed genes (LDSC-SEG) and multi-marker analysis of GenoMic annotation (MAGMA) were utilized to investigate single-nucleotide polymorphisms (SNP) enrichment in the tissue and cell-type levels. We then identified shared risk SNPs using cross-trait meta-analyses and Heritability Estimation from Summary Statistics (ρ-HESS). We further explore the potential functional genes for BMI and MS using summary-data-based Mendelian randomization (SMR).

**Result:** We found significantly positive genetic correlation and 18 novel shared genetic SNPs were identified in cross-trait meta-analyses. We found the causality of BMI on MS using Mendelian randomization, but slight inconsistent evidence for the causality of MS on BMI. We observed tissue-specific level SNP heritability enrichment for BMI in 9 tissues and MS in 4 tissues, and in cell-type-specific level SNP heritability enrichment 12 consistent cell types were identified for BMI and MS in brain, spleen, lung and whole blood.

**Conclusion:** Our study identifies the genetical correlation and shared risk SNPs between BMI and MS. These findings could provide new insights into the etiology of comorbidity and have implications for future therapeutic trials.

## Introduction

Obesity described by body mass index(BMI) is an increasing metabolic disorders in a large-scale age around the world^1,2^. The recent study has identified that obesity is the number one lifestyle related risk factor for premature death^3^. Observational studies have provided evidence that obesity is a significantly long-term risk^4^of suffering from multiple sclerosis (MS) in recent years^5,6^. Multiple sclerosis is a debilitating chronic demyelinating and neurodegenerative disease of the central nervous system (CNS)^7^. The acute and chronic disruption of white matter tracts and gray matter structure are contributing in the neurologic symptoms which may be incapacitating for patients and reducing patients quality of life and even leading disability^7–9^.

The epidemiological studies have revealed the intricate association between obesity and MS^10,11^. The BMI of MS patients were found have clear difference with healthy people and higher BMI in adolescence and childhood was considered as an environmental risk factor for MS ^12^,but whether MS patient have higher BMI was controversial ^13,14^. Mokry et al performed the first Mendelian randomization (MR) between elevated BMI and MS, providing positive evidence for a causal role for obesity in MS etiology^15^. A similar result was obtained by another MR analysis of a cohort involving 14,802 MS cases and 26,703 controls^16^. However, a large scale meta-analysis with 6,228 participants found that MS patients had lower BMI than healthy controls^17^. And a recent meta-analysis including 31 studies published between 1997 and 2020 indicated BMI does not present significant differences between patients with MS and healthy controls^18^. As a consequence, what roles of BMI play during the development of MS need further study ^19,20^.

In recent years, researchers have identified the common pathophysiological and immune elements for obesity and multiple sclerosis including inflammatory reaction^21,22^, the hormonal changes^23,24^ the interaction of the intestinal microbiome^25,26^ and the nutrition^27,28^, which are not mutually exclusive but may overlap^29^. In addition, during the clinical course, there are some drug interactions in patients with both MS and obesity^30^. According to an observational trail, obesity was proved to have an negative impact on IFNβ-treatment response in multiple sclerosis^31^. These findings suggest that the shared genetic background may exist between MS and BMI, whereas the magnitude of the genetic overlap required deep exploration^32,33^. Resolving these problems is benefit for making a clear understanding of the biological mechanisms and identify novel pharmacological targets for a precision medicine.

In this study, we conducted five complementary studies (**Fig. 1**) to investigate: (1) genetic correlation between BMI and MS using genome-wide association study (GWAS) summary statistics; (2) risk SNPs from Heritability Estimation from Summary Statistics(ρ-HESS) and cross-trait GWAS meta-analysis; (3) causality relationship between BMI and MS based on five Mendelian randomization methods; (4) SNP heritability enrichment in tissue levels and cell-type levels using GWAS data, Genotype-Tissue Expression (GTEx) dataset, and scRNA-seq dataset; and (5) putative functional genes for obesity and MS using Summary-data-based Mendelian Randomization(SMR).

**Figure1.**
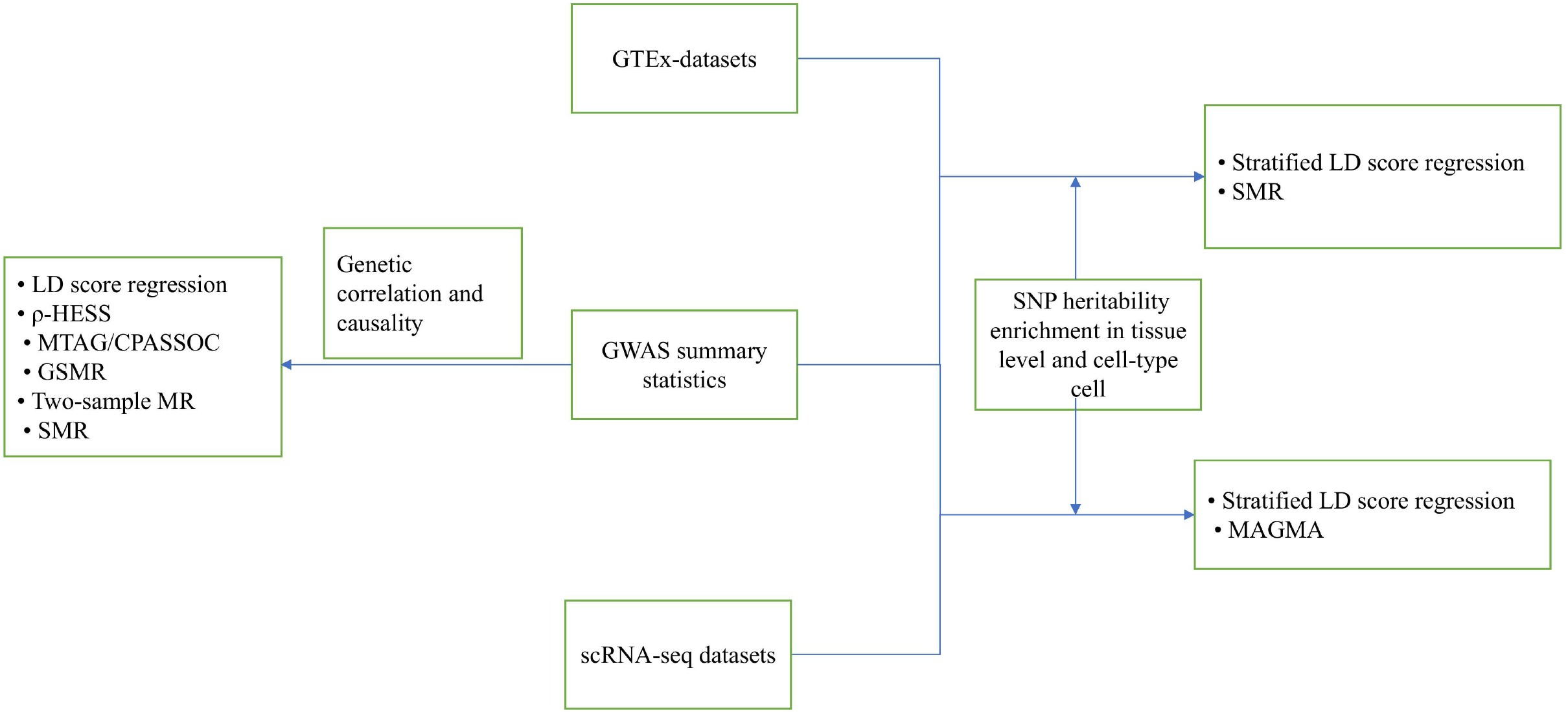
Overview of statistical analyses performed in the study. LD: linkage disequilibrium. GNOVA: Genetic covariance analyzer; ρ-HESS: Heritability Estimation from Summary Statistics. MTAG: Multi-Trait Analysis of GWAS. CPASSOC (Cross Phenotype Association). GSMR: Summary-data-based Mendelian Randomization. MR: Mendelian Randomization. GWAS: Genome-wide Association Study. scRNA-seq: single-cell RNA sequencing. MAGMA: Multi-marker Analysis of GenoMic Annotation. SMR: Summary-databased Mendelian randomisation.

## Methods

### Data samples

#### GWAS summary statistics

Effect estimates for SNPs associated with BMI were obtained from the Genetic Investigation of Anthropometric Traits (GIANT) consortium, which is a meta-analysis involving 2.4 million HapMap 2 (HM2) SNPs with available summary statistics in GIANT, 681 275 participants on average for BMI^35^. GWAS summary results for MS were derived from the International MS Genetics Consortium (IMSGC) meta-analysis of 15 datasets concluding 26,703 controls and 14,802 MS cases of European ancestry^34^. The publication described the collection of samples, quality control procedures, and imputation methods for each of the GWAS summary statistics^34,35^. GWAS protocols were approved by the appropriate ethics committees or institutional review boards, and all participants signed informed consent forms. The description of the studies is provided in **Supplementary Table S1**.

#### Bulk-tissue RNA sequencing gene expression data

In the subsequent LDSC-SEG and SMR analysis, we obtained bulk-tissue RNA-seq gene expression data from the Genotype-Tissue Expression (GTEx) project, which is a public data resource of gene expression in 53 non-diseased human primary tissues^36^. We download the GTEx v6p dataset which have been fixed^39^. We then choose the lite version of the GTEx V8 expression quantitative trait locus (eQTL) summary data (only SNPs with *P* < 1×10^−5^ are included). cis-eQTL summary statistics for whole blood were download for the downstream analysis from eQTLGen, a meta-analysis of 14,115 individuls^40^,

#### Single-cell RNA sequencing gene expression data

We obtained four single-cell RNA sequencing (scRNA-seq) resources from human lung^41^ (N□=□57,020 cells), spleen^41^ (N□=□94,257 cells) and whole blood from 10X Genomics Chromium and mouse brain^42^ (N= 133,509,876 cells). The “EWCE” R package^43^ were utilized to process the scRNA-seq data and convert mouse genes to human gene symbols.

### Statistical analyses

#### Heritability and Genetic Correlation

LD score regression (LDSC)^44^ (Python 2.7) is a useful method to estimate the genetic correlation for multiple traits or diseases. Based on the pre-computed LD scores of the 1000 Genomes projects which were calculated for SNPs in the HapMap 3 SNP set, we removed SNPs which did not match the reference panel (MAF ≤ 0.01 or INFO score ≤ 0.9) and reformatted new GWAS summary statistic^45^. We estimate single trait SNP heritability for BMI and MS using stratified linkage disequilibrium score regression (SLDSC) with the baseline-LD model^2^. According to recommendation, we set the parameter that the population prevalence and observed sample prevalence as 0.0003 and 0.63 separately to convert observed scale heritability to the liability-scale^46^. Then we performed bivariate LDSC^47^ without constraining the intercept to estimate *r*_*g*_ value, representing genetic correlations between MS and BMI and selected the suggestive (*P* < 0.05) genetic associations as the significant correlation^48^. Sensitivity analysis were conducted based on LDSC with the single-trait heritability intercept constrained. Because there was no sample overlap in our two traits, so we set all single-trait intercepts to 1 and all cross-trait intercepts to 0.

Genetic covariance analyzer (GNOVA)^49^ was supplemented to estimate the SNP-based heritability and genetic correlation between BMI and MS. GNOVA estimates genetic covariance based on all genetic variants shared between two GWAS summary statistics. Genetic correlation was then calculated relied on variant heritability and genetic covariance. Calculations were based on the 1000 Genomes Project European population-derived reference data using default parameters. In addition, sample overlap correction between two different sets of GWAS summary statistics was statistically calculated. Compared to LDSC, GNOVA provides higher estimation accuracy for genetic correlations and more powerful statistical inference^49^.

#### Identification of local genetic correlations using ρ-HESS

Heritability Estimation from Summary Statistics (ρ-HESS)^50^ is a method to estimate local SNP-heritability and genetic correlation from GWAS summary data. We estimated the local genetic correlations to examine whether BMI shared genetic correlation with MS at the local independent region in the genome using ρ-HESS (Python 2.7). There were 1699 potentially regions that were approximately LD-independent loci with average size of nearly 1.5Mb^51^. Then we calculated the local SNP heritability for two traits and genetic correlation between two traits using the 1000 Genomes project as reference provided on the HESS webpage^52^.

#### Cross-trait meta-analysis

To detect the shared risk SNPs in BMI and MS, we performed two cross-trait meta-analysis, including multi-trait analysis of GWAS (MTAG)^53^and cross-phenotype association test (CPASSOC)^54,55^. MTAG is a generalized meta-analysis method that enhances statistical power to estimates the genotypic and phenotypic variance-covariance matrices to generate trait-specific estimates for each SNP. SNPs were restricted with a minor allele frequency (MAF)□≥ 0.01 and sample size N ≥ (2/3) × 90^th^ percentile. MTAG adjusts for possible errors by bivariate LD score regression when sample overlap is present. MTAG is suitable when all variants have the same effect sizes on traits and generatestrait-specific association statistics. We calculated the upper bound for the false discovery rate (‘maxFDR’) to examine the assumptions on the equal variance-covariance. In addition, as a sensitivity analysis, CPASSOC integrates association evidence from multiple traits GWAS summary statistics, when variant is correlated to at least one trait. We utilized the SHet version to assume heterogeneous effects across traits. The SNP set is obtained from applying pairwise LD pruning with r^2^ = 0.2 using the software “PLINK”. We prioritize independent SNPs that were genome-wide significant (*P* < 5 × 10^−8^) in the cross-trait meta-analyses using both MTAG and CPASSOC and were in significant regions identified by ρ-HESS.

### MR analyses

To explore putative causal relationships between BMI and MS, using the R packages “TwoSample” and “GSMR” for the suggestive associations (*P* < 0.05). We undertook Mendelian Randomization analysis including main five MR methods, MR-Egger^56^, inverse variance weighting (IVW)^57^, weighted median^58^, weighted mode^59^ and Generalized Summary-data-based Mendelian Randomization (GSMR)^60^ with different assumptions about horizontal pleiotropy. Briefly, when there is one single genetic variant, the Wald ratio is a way to calculate the causative effect between the exposure and the outcome^48^. Using the meta-analysis approach, IVW analysis can estimate causal effects of two phenotype. It is a weighted average of the causal effects of genetic variants^57^.MR-Egger method further added a weighted linear regression of the gene–outcome coefficients for non-measured horizontal pleiotropy which allows for the presence of directional uncorrelated pleiotropy^56^. Pleiotropy test and Heterogeneity test were conducted by the MR-Egger intercept test^61^and Cochran’s Q statistic^62,63^, respectively, based on the “TwoSample” R package^64^. *P* < 0.05 is the level of statistical significance. For pleiotropy and outlier instrument detection, we choose the single SNP effect analysis ^65,66^ and MR-PRESSO analysis^61^.As for GSMR analysis, it assumes no correlation in pleiotropy but implements the HEIDI-outlier approach to test and exclude significant uncorrelated pleiotropy SNPs. For these five methods, we selected SNPs with genome-wide significance (*P* ≤ 5 × 10^−8^) of the ‘exposure’ trait as instrumental variables. Finally, all five methods were implemented bi-directional MR analyses.

### Linkage Disequilibrium Score Regression in specifically expressed genes (LDSC-SEG) analysis

We performed LDSC applied to specifically expressed genes (LDSC-SEG)^67^ to investigate whether SNP heritability for BMI and MS as evidence for trait-tissue relevance inference. The 1000 Genomes Phase 3 of European ancestry was utilized as a reference panel to calculate LD scores and SNPs only in HapMap 3 with MAF > 0.05 were included as input. Based on the baseline model and all gene sets, we ranked genes from the GTEx project by computed t-statistics reflecting critical tissue types and their specific expression in 53 tissues. We obtained the top 10% specifically expressed candidate genes with the highest t-statistic to estimate the significance of tissue type-specific SNP heritability enrichment. The coefficient *P*-values were calculated based on the regression coefficient Z-score, and Benjamini-Hochberg FDR-corrected *P*-value of <□5 × 10^−3^ was determined significance for enrichment tissues across the two traits.

### Cell type enrichment analyses using scRNA-seq datasets

We conducted Multi-marker Analysis of GenoMic Annotation (MAGMA)^68^ celltyping to evaluate whether gene-level genetic correlation between BMI and MS GWAS traits and cell type expression specificity. We using four scRNA-seq resources from human lung (N□=□57,020 cells), spleen (N□=□94,257 cells) and whole blood from 10X Genomics Chromium and mouse brain (N= 133,509,876 cells). Cell types across the four tissues were considered significant in MAGMA with *P*-value <0.05 after BH correction. The cell type specificity matrix for scRNA seq used in the MAGMA was calculated using Expression Weighted Cell type Enrichment, “EWCE” and “MAGMA_Celltyping” R package^69^.

### Summary-data-based Mendelian randomization

We conducted a SMR analysis^70^ to identify candidate risk genes of possibly causal effect and SNPs significant in cross-trait meta-analyses of BMI and MS. We used GWAS and eQTL data to detect the association between trait-associated SNPs and gene expression. The heterogeneity in dependent instrument (HEIDI) test was applied to distinguish linkage in the casual association. Genomic expression data from GTEx V8^71^ and cis-eQTL summary data from eQTLGen^40^ were used for the eQTL expression data of whole blood. As a default, we removed SNPs in strong LD with a r^2^ > 0.9 and with top associated eQTLs if MAF > 0.01. Expression probes with eQTL *P*□≤□5 × 10^−8^ were selected as top associated variants. All the SNPs were extracted by the genome-wide complex trait analysis (GCTA) software^72^, ensuring their independence. SMR significant probes were selected using Bonferroni-corrected thresholds for SMR *P*-values (0.05/number of probes) and HEIDI test *P*-value thresholds > 0.05 to indicate heterogeneity.

## Results

### Estimation of Genetic correlations between obesity and MS

We firstly calculate the liability-scale SNP heritability for BMI and MS through SLDSC^44^ with the baseline-LD model^73^. Consistent with the literature^33,74^, the liability-scale SNP heritability (without constrained intercept) was 4.6% for MS and 21% for BMI. We then used bivariate LDSC to estimate the genetic correlation (without constrained intercept) between BMI and MS (r_*g*_ = 0.0796, *P* = 0.0002). The intercept of genetic covariance was calculated at around 0.0021, indicating mild sample overlap between BMI and MS. After constraining the LDSC intercept on the assumption of no sample overlap, genetic correlation was slightly weaker (**Fig. 2)**. Moreover, all these estimates remained significant (*P* < 0.05, **Supplemental Table S1)**. To examine the robustness of our results, we performed GNOVA and identified a positive and consistent genetic association between BMI and MS with sample overlap corrected (*r*_g_ = 0.0647, *P*□= 3.36 ×□10^−5^), and the heritability estimates were 18.8% and 13.7% for the BMI and MS traits respectively.

**Figure2.**
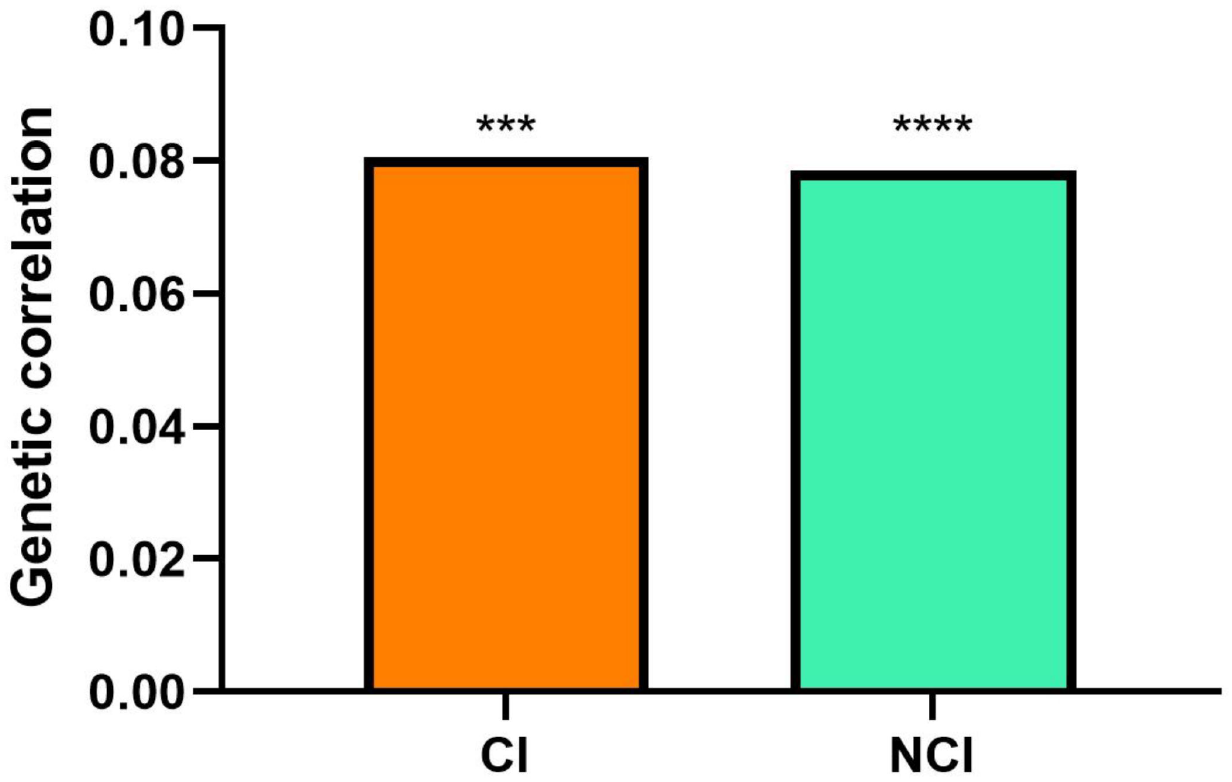
Summary of pairwise genetic correlations estimated using LDSC with and without constrained intercept. LDSC, Linkage disequilibrium score regression.

### Local genetic correlations between BMI and MS

Local genetic correlation was estimated for 1699 genomic partitions and 57 significant regions was identified (*P* < 0.05, **Supplemental Table S3)**. There was close agrrement in the average local genetic correlation in regions harbouring BMI-specific loci or MS-specific loci (**Fig. 3, Supplemental Fig.1**). We estimated the local single-trait SNP heritability for BMI (h^2^= 22.4%) and MS (h^2^= 23.4%) (**Supplemental Table S1**). Compared with bivariate LDSC, genome-wide local genetic correlations calculated by HESS between MS and BMI (*r*_g_ = 0.0428) were all largely consistent (**Supplemental Table S1**). These results suggest that across the whole genome, BMI and MS have potential correlation for sharing of genetic variation but not in specific genomic regions.

**Figure3.**
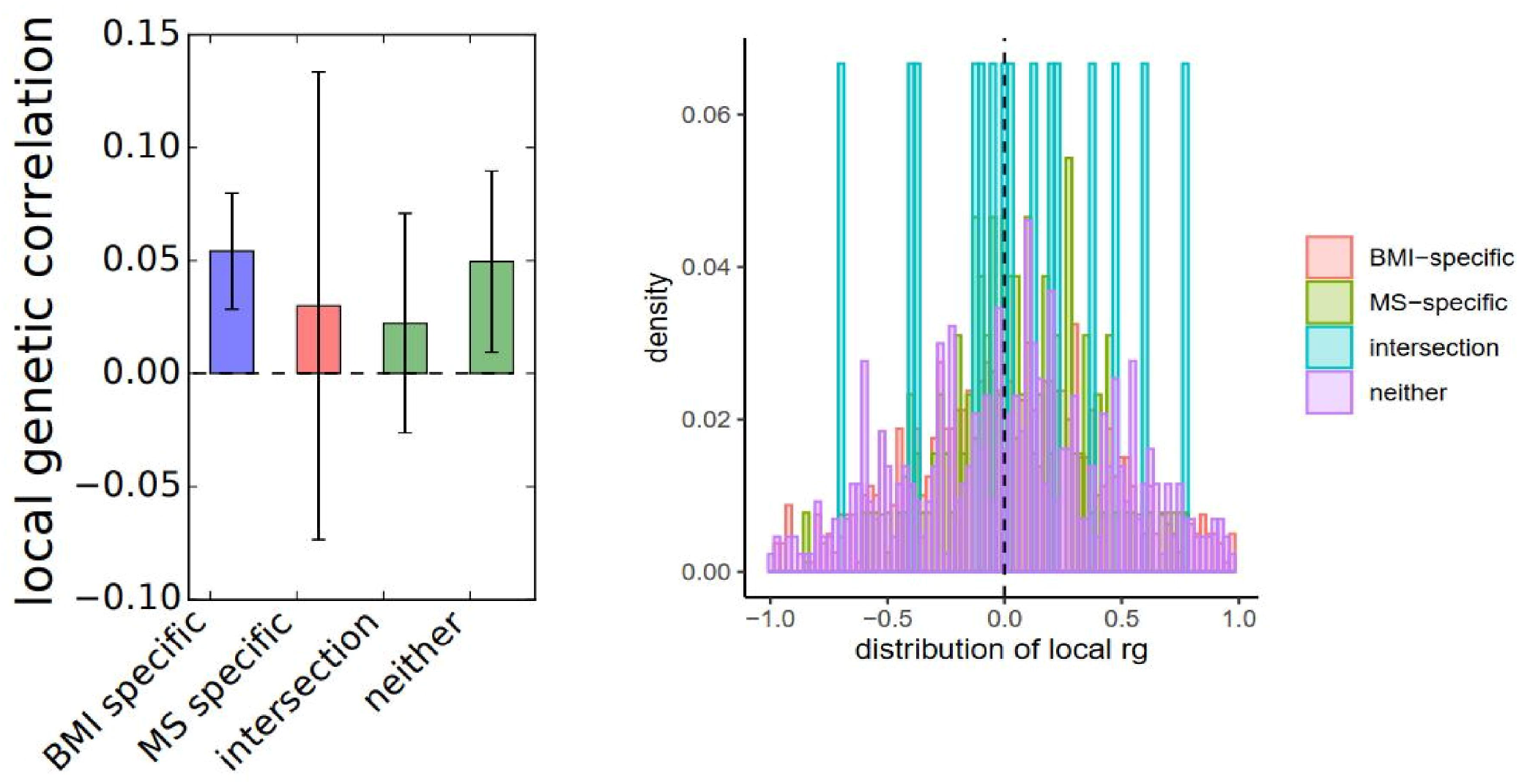
Summary of bi-directional MR analyses between BMI and MS. Purple: Generalized Summary-data-based Mendelian Randomization (GSMR); Pink: MR-Egger; Orange: inverse variance weighting (IVW); Sea blue: weighted median; Wathet: weighted mode. Error bars represent the 95% confidence intervals (CIs) for the associated MR point estimates; BMI, body mass index; MS, multiple sclerosis;

### Identification of Genomic risk SNPs for BMI and MS

Given the strong genetic relationships between BMI and MS, we conducted two cross-trait meta-analyses, MTAG and CPASSOC, to improve our power to identify genetic SNPs shared between traits. A total of 39 genome-wide significant SNPs (*P* < 5 × 10^−8^) were revealed in both MTAG and CPASSOC (**Supplemental Table S4**), 18 of them (rs11647753 rs11649612, rs12716972, rs12716974, rs2289292, rs3889624, rs4487979, rs4689871, rs2386589, rs2386593, rs8879260, rs8882, rs9986189, rs1944821, rs11667487, rs2382299, rs4888762, rs8112975) were not previously reported. The maxFDR values for MTAG analyses of BMI and MS were 1.4 × 10^−6^ and 1.8× 10^−2^ respectively. Furthermore, the MTAG results were highly consistent with those generated by CPASSOC, indicating that the MTAG results are reliable and that any bias in MTAG assumptions is likely to be negligible. Eventually, 22 risk SNPs are consistent significant in local genetic correlation examined by ρ-HESS.

### Evidence for causality between BMI and MS

We conducted bi-directional MR to explore the potentially causal effect and whether shared genetic background between BMI and MS was consistent with pleiotropy. We conducted various (N = 5) bi-directional MR methods to test the stability of relationships for a more stable result. We found evidence to support the causality of BMI on MS in five methods (*P* < 0.05) (**Fig. 4 Supplemental Fig.2**). In the reverse analyses, all these methods but GSMR identified negative causal effect of BMI on MS GSMR (**Supplemental Table S2**).

**Figure4.**
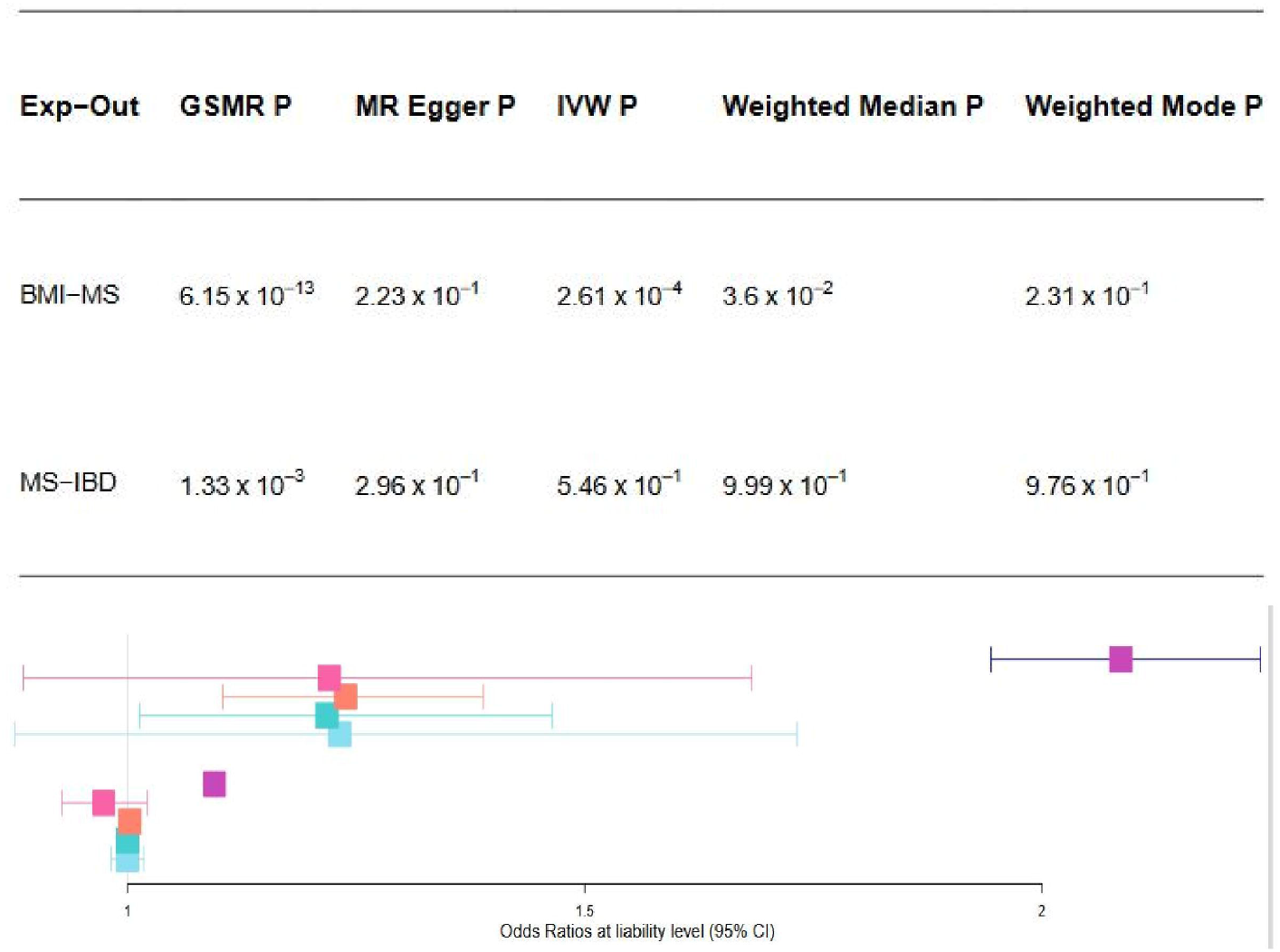
Local genetic correlations (rg) between BMI and MS. (A) Average local rg estimates for two traits in regions harbouring disease-specific risk variants (*P* < 5 ×10^−6^), regions harbouring shared risk variants (“intersection”) and all other regions (“neither”). Local genetic correlations with estimates less than −1 or greater than 1 were forced to −1 or 1, respectively. Error bars represent the 95% confidence intervals (CIs), calculated using a jackknife method. (B) density distribution of local rg estimates for two traits in disease-specific regions (red, green), intersection regions (blue) and other (purple) regions. BMI, body mass index; MS, multiple sclerosis;

### Tissue-level SNP heritability enrichment in BMI and MS

We conducted LDSC-SEG method to identify specific tissues in which genes with increased expression are enriched in SNPs, using publicly available GWAS data and genotype tissue expression data from GTEx. After adjusting for the baseline model, we identified FDR-significant (*P*□<□5 × 10^−3^) SNP heritability enrichment for BMI across 9 tissues **(Fig. 5A, Supplementary TableS5**), including frontal cortex, anterior cingulate cortex, nucleus accumbens, putamen, caudate, hypothalamus, cerebellar hemisphere, cerebellum and cortex, particularly for central nervous system (CNS)-related tisWsues. For MS, a total of 4 tissues were significant enrichment, including spleen, Epstein-Barr virus (EBV)-transformed lymphoblastoid cell lines (LCLs), lung and whole blood, particularly for blood and immune-related tissues (**Fig. 5B, Supplementary TableS5**).

**Figure5.**
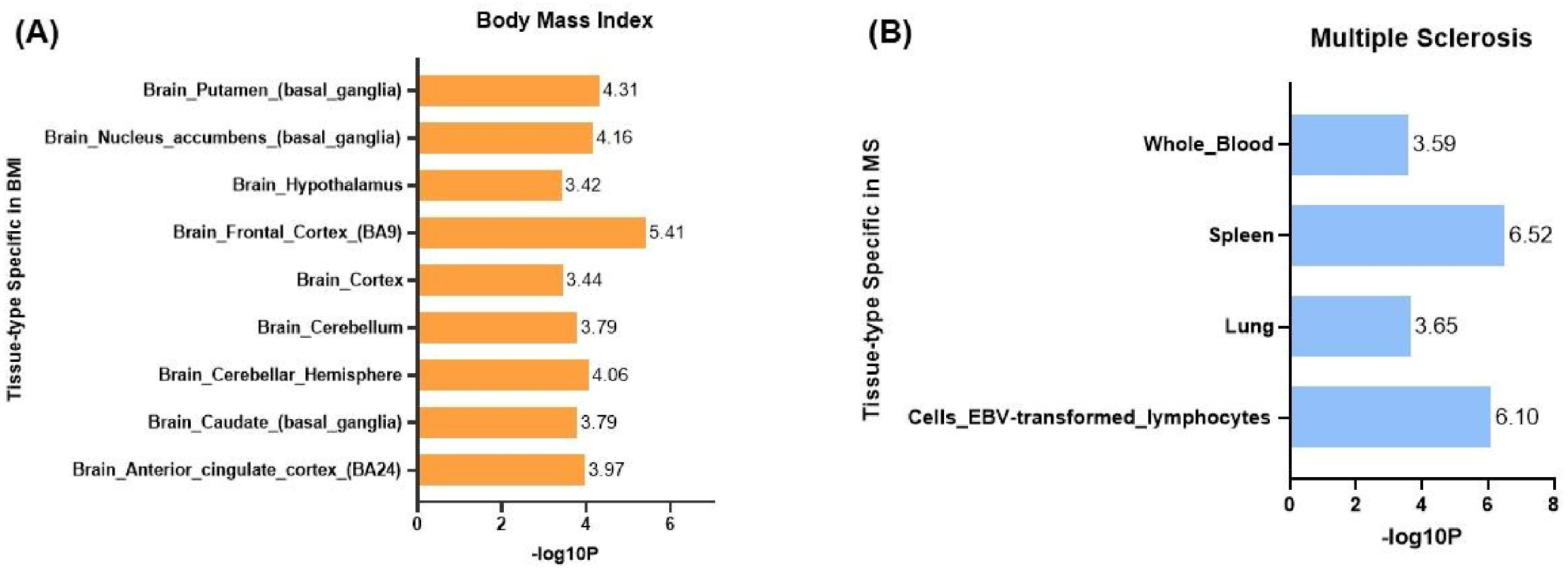
Tissue type-specific enrichment of SNP heritability for BMI and MS estimated using LDSC-SEG. (A) The heritability enrichment of tissues in BMI; (B) The heritability enrichment of tissues in MS; The x axis displays negative log10 *P*-values of coefficient Z-scores for each individual test. SNP, single nucleotide polymorphism; BMI, body mass index; MS, multiple sclerosis; LDSC-SEG: linkage disequilibrium score regression applied to specifically expressed genes.

### Cell-level SNP heritability enrichment in BMI and MS

We utilized publicly available scRNA-seq datasets of four tissues enriched in LDSC-SEG, including brain, spleen, lung and whole blood to evaluated the gene-level genetic association with cell type expression specificity for BMI and MS. In the lung dataset, we found both significant enrichment at *P* < 0.05 for BMI and MS in mature B cells, naive B cells, mast cells, natural killer cells (NK), dividing NK cells, CD4+ T cells, CD8+cytotoxic T lymphocytes, dividing T cells, regulatory T cells, activated dendritic (DC) cells, plasmacytoid DC cells and Monocytes. We observed a significant enrichment across NK_FCGR3Apos in spleen tissue. Cells in brain and in peripheral blood mononuclear cells (PBMC) tissues are not co-enriched in BMI and MS traits. The enriched cells in four tissues for BMI and MS were listed separately (**Supplemental Fig.3-6)**.

### Identification of shared functional genes for BMI and MS

We applied SMR to infer causality and identify the putative functional genes for BMI and MS, by jointly analyzing GWAS summary data and whole blood eQTL summary data from eQTLGen and GTEx. We identified 10 genome-wide significant associations (*P*□<□5.28 × 10^−7^), of which only one gene, the Gametogenetin-binding protein (*GGNBP2*), shared between BMI (*P*_SMR_□=□3.69 × 10^−19^, *P*_HEIDI□0_=□0.11, topSNP: rs11263770) and MS □ (*P*_SMR_□□=□2.41 × 10^−7^, *P*_HEIDI□0_=□0.98, topSNP: rs11650008) and passed the HEIDI-outlier □ test in cis_eQTL data (**Supplementary TableS6**). More importantly, the *GGNBP2* was also identified as one of the genetically shared variants in previous cross-trait meta-analysis phenotypes between BMI and MS.

## Discussion

We present the first assessment (to our knowledge) of overlapping polygenic architecture of BMI with MS by combining large-scale GWAS summary data, GTEx datasets and scRNA-seq datasets. Our results provide new insights into the molecular genetic mechanism for comorbidity and may contribute to the disease prediction and clinical treatment.

Our findings provided the evidence that there existed a significantly strong genetic correlation between BMI and MS, suggesting that genetic factors play an important role in comorbidity of obesity and MS. Analyzing local genetic correlation using ρ-HESS, the regional *r*_g_ estimates were close to global *r*_g_ estimates based on LDSC and GNOVA, suggesting pleiotropic effects on the two traits exist in numerous genetic variants across the genome^34^. Notwithstanding that multiple factors may lead to disease comorbidity^11,75^, our results support the risk of genetic factors.

From the cross-trait meta-analysis, 39 genetic SNPs identified in both MTAG and CPASSOC analysis, in which 22 SNPs located in the ρ-HESS estimated significant genomic regions. The novel SNPs we identified may be involved in regulating a common pathway shared between BMI and MS, advancing the understanding of the causes of risk for both traits. We used MTAG and CPASSOC, two completely independent statistical analysis methods, not only to avoid the possible bias caused by potential sample overlap but also to verify the risk SNPS obtained from MTAG with CPASSOC. Results showed that the SNPs identified by MTAG were all consistently significant in the CPASSOC analysis results, which adequately improved the reliability of our results.

We conducted Mendelian randomization studies using the latest GWAS summary data. A consistent causality of BMI on MS was inferred in all MR methods, showing no difference on previous study^15,76^. We performed several sensitivity analyses which provided little evidence of genetic pleiotropy, adding further robustness to the findings. It may owing to the different criteria for outliers so that bi-directional GSMR analysis inferred a causalty of MS on BMI but other methods inferred no effect of MS on BMI, which suggesting the significance of utilizing different methods in MR analyses^34^. The establishment of more accurately causal relationship requires larger and more powerful GWAS for BMI and MS.

In this study, we comprehensively investigated functional enrichment for gene expression in multiple tissues and cells by applying LDSC-SEG using the GTEx gene expression datasets. We identified 9 tissues, mainly brain, with significant SNP heritability enrichment for BMI. Growing evidence also suggests that susceptibility to obesity is distributed across multiple brain regions and strongly associated with structural abnormalities^77–81^. The enrichment results we obtained in MS were mainly reflected in immune tissues, including spleen and LCLs, suggesting the involvement of local immune responses in the process of MS. This is consistent with previous literature demonstrating a strong relationship between MS and immune dysregulation^82^. EBV-infected B cells and plasma cells accumulate in meningeal immune cell collections that may contribute to the progressive development of MS^83^. In the center of the immune response in MS is the CD4+ T cell^84^. The lung could contribute to activate and transform autoreactive T cells to a migratory mode to enter the CNS and induce autoimmune disease including MS^85^. Different immune pathways drive MS disease relapse and progression, and our approach may more specifically understand MS as a disease of certain key tissue and cellular processes. Further experimental validation of the enriched tissues and cells for diseases is still required.

Notably, we identified enrichments in mature B cells, naive B cells, mast cells, natural killer cells (NK), dividing NK cells, CD4+ T cells, CD8+cytotoxic T lymphocytes, dividing T cells, regulatory T cells, activated dendritic (DC) cells, plasmacytoid DC cells and Monocytes in lung and NK cells in spleen for both BMI and MS. Several cells-related genes. the heritability enrichment in CD8 cytotoxic T cells in lung and spleen for both BMI and MS, suggesting a possible role for these cell types in the comorbidity of obesity and MS. MS is an immunologically heterogeneous disorder, and CD8 + T cells predominate in MS lesions. B cell depletion may reduce the proinflammatory cytokines produced by B cells, CD4+ and CD8 + T cells which can effectively reduce MS relapses^86,87^. NK and DC cells control T cell activation in CNS autoimmunity, and reduced the risk of MS^88,89^. Mast cells participate in the pathogenesis of MS by promoting angiogenesis^90^. Our results may gain insights into the pathogenesis of comorbid MS or IBD and will be instrumental for further studies to develop new targeted therapies in specific tissues and cell types.

In addition to cross-trait meta-analysis, we also used large-scale cis_eQTL and whole-blood GTEx data to test whether BMI-MS association can be mediated by shared risk genes. Using SMR and HEIDI, we discovered one functional gene, *GGNBP2*, which might serve as a potential links between the two traits. Prior studies have documented the biological relationship between *GGNBP2* and BMI. *GGNBP2* is a tumor suppressor gene involved in several kinds of cancers, such as glioma^91^, breast cancer^92^ and prostate cancer^93^. Previous study has identified that *GGNBP2* is a shared genetic loci for ALS and obesity related traits, and associated with BMI and waist-hip ratio^94^. We first discovered the biological relationship between *GGNBP2* and the two traits. Further functional and other researches need to be undertaken to systematically investigate the biological mechanisms of the novel gene *GGNBP2* on obesity and MS risk.

Our study also has limitations. Firstly, we choose the GWAS data from GIANT and IMSGC which both used samples from the WTCCC cohort. Therefore, we were not sure if there exists the possibility of sample overlap, which might bias our results. However, given the sample size employed, this effect would likely be small since the WTCCC comprised ∼2.5% of the overall GIANT consortium^15^. Secondly, this study was restricted to individuals of European descent to prevent population stratification and therefore our findings have limited generalizability to other ancestral populations. Thirdly, although enrichment analysis revealed the potential overlapping genetic components but how the shared biological pathways work warrants further research.

## Conclusion

In summary, we found significant genetic correlation and identified novel shared risk SNPs between BMI and MS. We confirmed the causality of BMI on MS but reverse causality was inconclusive. We further explored SNP heritability enrichment for BMI and MS and found a putative functional gene *GGNBP2*.

These findings could provide novel insights into the genetic overlap between obesity and MS and contribute to better understand the etiology of comorbidity and may keep instrumental in the disease prediction and clinical treatment.

## Data Availability

All data produced in the present work are contained in the manuscript

## Data source

GWAS summary statistics for MS are available by application from: https://imsgc.net/? page_id=31.

GWAS summary statistics for MS are available by application from: http://portals.broadinstitute.org/collaboration/giant/index.php/GIANT_consortium_data_files. The eQTL summary data for eQTLGen and GTEx : https://www.eqtlgen.org/cis-eqtls.html http://yanglab.westlake.edu.cn/software/smr/#eQTLsummarydata. scRNA-seq data:

Whole blood: https://www.10xgenomics.com/resources/datasets;

Lung and Spleen: https://doi.org/10.1186/s13059-019-1906-x;

Brain: https://doi.org/10.1016/j.cell.2018.06.021;

Small Intestinal Epithelium: https://singlecell.broadinstitute.org/single_cell/study/SCP44/small-intestinal-epithelium

## Code availability

LDSC: https://github.com/bulik/ldsc.

PLINK: https://www.cog-genomics.org/plink/1.9.

MTAG: https://github.com/JonJala/mtag.

CPASSOC: http://hal.case.edu/~xxz10/zhuweb/.

GSMR: http://cnsgenomics.com/software/gsmr/.

TwoSampleMR:https://mrcieu.github.io/TwoSampleMR/.

SMR:https://cnsgenomics.com/software/smr/#Overview.

LDSC-SEG: https://github.com/bulik/ldsc/wiki/Cell-type-specific-analyses

MAGMA Celltyping: https://neurogenomics.github.io/MAGMA_Celltyping

## Author contributions

FWL, QZL, WHS and HC designed, revised and supervised the study; RJZ, RJ, WTH and JXW analyzed and organized the data; LJZ, YYM, YJW and MJM generated the figures and tables; RJZ, RJ, WTH and HC wrote this manuscript; All authors reviewed and approved the final manuscript.

## Funding

This work was funded by the National Natural Science Foundation of China (82171698, 82170561, 81300279, and 81741067), the Natural Science Foundation for Distinguished Young Scholars of Guangdong Province (2021B1515020003), Natural Science Foundation of Guangdong Province (2022A1515012081), the Climbing Program of Introduced Talents and High-level Hospital Construction Project of Guangdong Provincial People’s Hospital (DFJH201803, KJ012019099, KJ012021143, and KY012021183).

## Acknowledgements

We thank IMSGC and GIANT for providing access to their GWAS summary data. We gratefully acknowledge the contributions from public available databases.

## Conflict of interest statement

The authors declare that the research was conducted in the absence of any commercial or financial relationships that could be construed as a potential conflict of interest.

## Notes

### Competing Interest Statement

The authors have declared no competing interest.

### Author Declarations

GWAS summary statistics for MS are available by application from: https://imsgc.net/?page_id=31. GWAS summary statistics for MS are available by application from: http://portals.broadinstitute.org/collaboration/giant/index.php/GIANT_consortium_data_files. The eQTL summary data for eQTLGen and GTEx : https://www.eqtlgen.org/cis-eqtls.html http://yanglab.westlake.edu.cn/software/smr/#eQTLsummarydata https://www.eqtlgen.org/cis-eqtls.html http://yanglab.westlake.edu.cn/software/smr/#eQTLsummarydata. scRNA-seq data: Whole blood: https://www.10xgenomics.com/resources/datasets; Lung and Spleen: https://doi.org/10.1186/s13059-019-1906-x; Brain: https://doi.org/10.1016/j.cell.2018.06.021; Small Intestinal Epithelium: https://singlecell.broadinstitute.org/single_cell/study/SCP44/small-intestinal-epithelium

## References

1. Chen, X.-W., Ding, G., Xu, L. & Li, P. A glimpse at the metabolic research in China. Cell Metab 33, 2122–2125 (2021).

2. Hu, K. & Staiano, A. E. Trends in Obesity Prevalence Among Children and Adolescents Aged 2 to 19 Years in the US From 2011 to 2020. JAMA Pediatr 176, 1037–1039 (2022).

3. Chooi, Y. C., Ding, C. & Magkos, F. The epidemiology of obesity. Metabolism 92, 6–10 (2019).

4. Høglund, R. A. A. et al. Association of Body Mass Index in Adolescence and Young Adulthood and Long-term Risk of Multiple Sclerosis: A Population-Based Study. Neurology 97, e2253–e2261 (2021).

5. Munger, K. L. et al. Childhood body mass index and multiple sclerosis risk: a long-term cohort study. Mult Scler 19, 1323–1329 (2013).

6. Hedström, A. K., Olsson, T. & Alfredsson, L. High body mass index before age 20 is associated with increased risk for multiple sclerosis in both men and women. Mult Scler 18, 1334–1336 (2012).

7. Olek, M. J. Multiple Sclerosis. Ann Intern Med 174, ITC81–ITC96 (2021).

8. Kobelt, G. et al. New insights into the burden and costs of multiple sclerosis in Europe. Mult Scler 23, 1123–1136 (2017).

9. Ness, N.-H. et al. Differentiating societal costs of disability worsening in multiple sclerosis. J Neurol 267, 1035–1042 (2020).

10. Kl, M. Childhood obesity is a risk factor for multiple sclerosis. Multiple sclerosis (Houndmills, Basingstoke, England) 19, (2013).

11. Schreiner, T.-G. & Genes, T.-M. Obesity and Multiple Sclerosis-A Multifaceted Association. J Clin Med 10, 2689 (2021).

12. Marrie, R. A. Environmental risk factors in multiple sclerosis aetiology. Lancet Neurol 3, 709–718 (2004).

13. Munger, K. L., Chitnis, T. & Ascherio, A. Body size and risk of MS in two cohorts of US women. Neurology 73, 1543–1550 (2009).

14. Bhargava, P. et al. Multiple sclerosis patients have a diminished serologic response to vitamin D supplementation compared to healthy controls. Mult Scler 22, 753–760 (2016).

15. Mokry, L. E. et al. Obesity and Multiple Sclerosis: A Mendelian Randomization Study. PLoS Med 13, e1002053 (2016).

16. Harroud, A. et al. The relative contributions of obesity, vitamin D, leptin, and adiponectin to multiple sclerosis risk: A Mendelian randomization mediation analysis. Mult Scler 27, 1994–2000 (2021).

17. Dardiotis, E. et al. Body mass index in patients with Multiple Sclerosis: a meta-analysis. Neurol Res 41, 836–846 (2019).

18. Castro, L. J. J. O., Pereira, A. & Ribeiro, N. The Body mass index in patients with Multiple Sclerosis: A meta-analysis. Journal of Statistics on Health Decision 3, 99–104 (2021).

19. Hedström, A. K., Olsson, T. & Alfredsson, L. Body mass index during adolescence, rather than childhood, is critical in determining MS risk. Mult Scler 22, 878–883 (2016).

20. Langer-Gould, A., Brara, S. M., Beaber, B. E. & Koebnick, C. Childhood obesity and risk of pediatric multiple sclerosis and clinically isolated syndrome. Neurology 80, 548–552 (2013).

21. Giovannoni, G. The neurodegenerative prodrome in multiple sclerosis. Lancet Neurol 16, 413–414 (2017).

22. Stampanoni Bassi, M. et al. Obesity worsens central inflammation and disability in multiple sclerosis. Mult Scler 26, 1237–1246 (2020).

23. Dragano, N. R. V., Haddad-Tovolli, R. & Velloso, L. A. Leptin, Neuroinflammation and Obesity. Front Horm Res 48, 84–96 (2017).

24. Izquierdo, A. G., Crujeiras, A. B., Casanueva, F. F. & Carreira, M. C. Leptin, Obesity, and Leptin Resistance: Where Are We 25 Years Later? Nutrients 11, E2704 (2019).

25. Dalile, B., Van Oudenhove, L., Vervliet, B. & Verbeke, K. The role of short-chain fatty acids in microbiota-gut-brain communication. Nat Rev Gastroenterol Hepatol 16, 461–478 (2019).

26. Buscarinu, M. C. et al. Altered intestinal permeability in patients with relapsing-remitting multiple sclerosis: A pilot study. Mult Scler 23, 442–446 (2017).

27. Parks, N. E., Jackson-Tarlton, C. S., Vacchi, L., Merdad, R. & Johnston, B. C. Dietary interventions for multiple sclerosis-related outcomes. Cochrane Database Syst Rev 5, CD004192 (2020).

28. Sanchez, J. M. S., DePaula-Silva, A. B., Libbey, J. E. & Fujinami, R. S. Role of diet in regulating the gut microbiota and multiple sclerosis. Clin Immunol 235, 108379 (2022).

29. Wanleenuwat, P. & Iwanowski, P. Role of B cells and antibodies in multiple sclerosis. Mult Scler Relat Disord 36, 101416 (2019).

30. Manni, A. et al. Lymphocyte Count and Body Mass Index as Biomarkers of Early Treatment Response in a Multiple Sclerosis Dimethyl Fumarate-Treated Cohort. Front Immunol 10, 1343 (2019).

31. Kvistad, S. S. et al. Body mass index influence interferon-beta treatment response in multiple sclerosis. Journal of Neuroimmunology 288, 92–97 (2015).

32. Wen, W. et al. Meta-analysis identifies common variants associated with body mass index in east Asians. Nat Genet 44, 307–311 (2012).

33. International Multiple Sclerosis Genetics Consortium. Multiple sclerosis genomic map implicates peripheral immune cells and microglia in susceptibility. Science 365, eaav7188 (2019).

34. Yang, Y. et al. Investigating the shared genetic architecture between multiple sclerosis and inflammatory bowel diseases. Nat Commun 12, 5641 (2021).

35. Yengo, L. et al. Meta-analysis of genome-wide association studies for height and body mass index in ∼700000 individuals of European ancestry. Hum Mol Genet 27, 3641–3649 (2018).

36. GTEx Consortium et al. Genetic effects on gene expression across human tissues. Nature 550, 204–213 (2017).

37. Pers, T. H. et al. Biological interpretation of genome-wide association studies using predicted gene functions. Nat Commun 6, 5890 (2015).

38. Fehrmann, R. S. N. et al. Gene expression analysis identifies global gene dosage sensitivity in cancer. Nat Genet 47, 115–125 (2015).

39. Finucane, H. K. et al. Heritability enrichment of specifically expressed genes identifies disease-relevant tissues and cell types. Nat Genet 50, 621–629 (2018).

40. Võsa, U. et al. Unraveling the polygenic architecture of complex traits using blood eQTL metaanalysis. 447367 Preprint at https://doi.org/10.1101/447367 (2018).

41. Madissoon, E. et al. scRNA-seq assessment of the human lung, spleen, and esophagus tissue stability after cold preservation. Genome Biol 21, 1 (2019).

42. Zeisel, A. et al. Molecular Architecture of the Mouse Nervous System. Cell 174, 999-1014.e22 (2018).

43. Skene, N. G. & Grant, S. G. N. Identification of Vulnerable Cell Types in Major Brain Disorders Using Single Cell Transcriptomes and Expression Weighted Cell Type Enrichment. Front Neurosci 10, 16 (2016).

44. Hk, F. et al. Partitioning heritability by functional annotation using genome-wide association summary statistics. Nature genetics 47, (2015).

45. 1000 Genomes Project Consortium et al. A global reference for human genetic variation. Nature 526, 68–74 (2015).

46. Wallin, M. T. et al. The prevalence of MS in the United States: A population-based estimate using health claims data. Neurology 92, e1029–e1040 (2019).

47. Bulik-Sullivan, B. et al. An atlas of genetic correlations across human diseases and traits. Nat Genet 47, 1236–1241 (2015).

48. Yao, Y. et al. An atlas of genetic correlations between gestational age and common psychiatric disorders. Autism Res 15, 1008–1017 (2022).

49. Lu, Q. et al. A Powerful Approach to Estimating Annotation-Stratified Genetic Covariance via GWAS Summary Statistics. Am J Hum Genet 101, 939–964 (2017).

50. H, S., N, M., S, S. & B, P. Local Genetic Correlation Gives Insights into the Shared Genetic Architecture of Complex Traits. American journal of human genetics 101, (2017).

51. Berisa, T. & Pickrell, J. K. Approximately independent linkage disequilibrium blocks in human populations. Bioinformatics 32, 283–285 (2016).

52. Stolp Andersen, M. et al. Dissecting the limited genetic overlap of Parkinson’s and Alzheimer’s disease. Ann Clin Transl Neurol 9, 1289–1295 (2022).

53. 23 andMe Research Team et al. Multi-trait analysis of genome-wide association summary statistics using MTAG. Nat Genet 50, 229–237 (2018).

54. Zhu, X. et al. Meta-analysis of correlated traits via summary statistics from GWASs with an application in hypertension. Am J Hum Genet 96, 21–36 (2015).

55. Li, X. & Zhu, X. Cross-Phenotype Association Analysis Using Summary Statistics from GWAS. Methods Mol Biol 1666, 455–467 (2017).

56. Burgess, S. & Thompson, S. G. Interpreting findings from Mendelian randomization using the MR-Egger method. Eur J Epidemiol 32, 377–389 (2017).

57. Burgess, S., Butterworth, A. & Thompson, S. G. Mendelian randomization analysis with multiple genetic variants using summarized data. Genet Epidemiol 37, 658–665 (2013).

58. Bowden, J., Davey Smith, G., Haycock, P. C. & Burgess, S. Consistent Estimation in Mendelian Randomization with Some Invalid Instruments Using a Weighted Median Estimator. Genet Epidemiol 40, 304–314 (2016).

59. Hartwig, F. P., Davey Smith, G. & Bowden, J. Robust inference in summary data Mendelian randomization via the zero modal pleiotropy assumption. Int J Epidemiol 46, 1985–1998 (2017).

60. Zhu, Z. et al. Causal associations between risk factors and common diseases inferred from GWAS summary data. Nat Commun 9, 224 (2018).

61. Verbanck, M., Chen, C.-Y., Neale, B. & Do, R. Detection of widespread horizontal pleiotropy in causal relationships inferred from Mendelian randomization between complex traits and diseases. Nat Genet 50, 693–698 (2018).

62. Liu, G. et al. PICALM gene rs3851179 polymorphism contributes to Alzheimer’s disease in an Asian population. Neuromolecular Med 15, 384–388 (2013).

63. Greco M F. D., Minelli, C., Sheehan, N. A. & Thompson, J. R. Detecting pleiotropy in Mendelian randomisation studies with summary data and a continuous outcome. Stat Med 34, 2926–2940 (2015).

64. Yavorska, O. O. & Burgess, S. MendelianRandomization: an R package for performing Mendelian randomization analyses using summarized data. Int J Epidemiol 46, 1734–1739 (2017).

65. Wang, R. Mendelian randomization study updates the effect of 25-hydroxyvitamin D levels on the risk of multiple sclerosis. J Transl Med 20, 3 (2022).

66. Noyce, A. J. et al. Estimating the causal influence of body mass index on risk of Parkinson disease: A Mendelian randomisation study. PLoS Med 14, e1002314 (2017).

67. Gazal, S., Marquez-Luna, C., Finucane, H. K. & Price, A. L. Reconciling S-LDSC and LDAK functional enrichment estimates. Nat Genet 51, 1202–1204 (2019).

68. de Leeuw, C. A., Mooij, J. M., Heskes, T. & Posthuma, D. MAGMA: generalized gene-set analysis of GWAS data. PLoS Comput Biol 11, e1004219 (2015).

69. Skene, N. G. et al. Genetic identification of brain cell types underlying schizophrenia. Nat Genet 50, 825–833 (2018).

70. Zhu, Z. et al. Integration of summary data from GWAS and eQTL studies predicts complex trait gene targets. Nat Genet 48, 481–487 (2016).

71. GTEx Consortium. The GTEx Consortium atlas of genetic regulatory effects across human tissues. Science 369, 1318–1330 (2020).

72. Yang, J., Lee, S. H., Goddard, M. E. & Visscher, P. M. GCTA: a tool for genome-wide complex trait analysis. Am J Hum Genet 88, 76–82 (2011).

73. Gazal, S. et al. Linkage disequilibrium-dependent architecture of human complex traits shows action of negative selection. Nat Genet 49, 1421–1427 (2017).

74. Wang, R., Snieder, H. & Hartman, C. A. Familial co-aggregation and shared heritability between depression, anxiety, obesity and substance use. Transl Psychiatry 12, 1–8 (2022).

75. Alfredsson, L. & Olsson, T. Lifestyle and Environmental Factors in Multiple Sclerosis. Cold Spring Harb Perspect Med 9, a028944 (2019).

76. Gianfrancesco, M. A. et al. Causal Effect of Genetic Variants Associated With Body Mass Index on Multiple Sclerosis Susceptibility. Am J Epidemiol 185, 162–171 (2017).

77. Willeumier, K. C., Taylor, D. V. & Amen, D. G. Elevated BMI is associated with decreased blood flow in the prefrontal cortex using SPECT imaging in healthy adults. Obesity (Silver Spring) 19, 1095–1097 (2011).

78. Opel, N. et al. Brain structural abnormalities in obesity: relation to age, genetic risk, and common psychiatric disorders□: Evidence through univariate and multivariate mega-analysis including 6420 participants from the ENIGMA MDD working group. Mol Psychiatry 26, 4839–4852 (2021).

79. Preston, C. & Ehrsson, H. H. Illusory Obesity Triggers Body Dissatisfaction Responses in the Insula and Anterior Cingulate Cortex. Cereb Cortex 26, 4450–4460 (2016).

80. Green, E., Jacobson, A., Haase, L. & Murphy, C. Reduced nucleus accumbens and caudate nucleus activation to a pleasant taste is associated with obesity in older adults. Brain Res 1386, 109–117 (2011).

81. Kurth, F. et al. Relationships between gray matter, body mass index, and waist circumference in healthy adults. Hum Brain Mapp 34, 1737–1746 (2013).

82. Dendrou, C. A., Fugger, L. & Friese, M. A. Immunopathology of multiple sclerosis. Nat Rev Immunol 15, 545–558 (2015).

83. Balfour, H. H., Schmeling, D. O. & Grimm-Geris, J. M. The promise of a prophylactic Epstein-Barr virus vaccine. Pediatr Res 87, 345–352 (2020).

84. International Multiple Sclerosis Genetics Consortium et al. Genetic risk and a primary role for cell-mediated immune mechanisms in multiple sclerosis. Nature 476, 214–219 (2011).

85. Odoardi, F. et al. T cells become licensed in the lung to enter the central nervous system. Nature 488, 675–679 (2012).

86. Comi, G. et al. Role of B Cells in Multiple Sclerosis and Related Disorders. Ann Neurol 89, 13–23 (2021).

87. Jelcic, I. et al. Memory B Cells Activate Brain-Homing, Autoreactive CD4+ T Cells in Multiple Sclerosis. Cell 175, 85-100.e23 (2018).

88. Piacente, F. et al. Neuroprotective Potential of Dendritic Cells and Sirtuins in Multiple Sclerosis. Int J Mol Sci 23, 4352 (2022).

89. Gross, C. C. et al. Regulatory Functions of Natural Killer Cells in Multiple Sclerosis. Front Immunol 7, 606 (2016).

90. Ribatti, D., Tamma, R. & Annese, T. Mast cells and angiogenesis in multiple sclerosis. Inflamm Res 69, 1103–1110 (2020).

91. Zhan, A. et al. GGNBP2 Suppresses the Proliferation, Invasion, and Migration of Human Glioma Cells. Oncol Res 25, 831–842 (2017).

92. Lan, Z.-J. et al. GGNBP2 acts as a tumor suppressor by inhibiting estrogen receptor α activity in breast cancer cells. Breast Cancer Res Treat 158, 263–276 (2016).

93. Yang, Z., Wang, Y. & Ma, L. Effects of gametogenetin-binding protein 2 on proliferation, invasion and migration of prostate cancer PC-3 cells. Andrologia 52, e13488 (2020).

94. Li, C., Ou, R., Wei, Q. & Shang, H. Shared genetic links between amyotrophic lateral sclerosis and obesity-related traits: a genome-wide association study. Neurobiol Aging 102, 211.e1-211.e9 (2021).

