## Supplementary figures and images for "Investigating shared genetic architecture between obesity and multiple sclerosis"

### Supplementary Figure S1

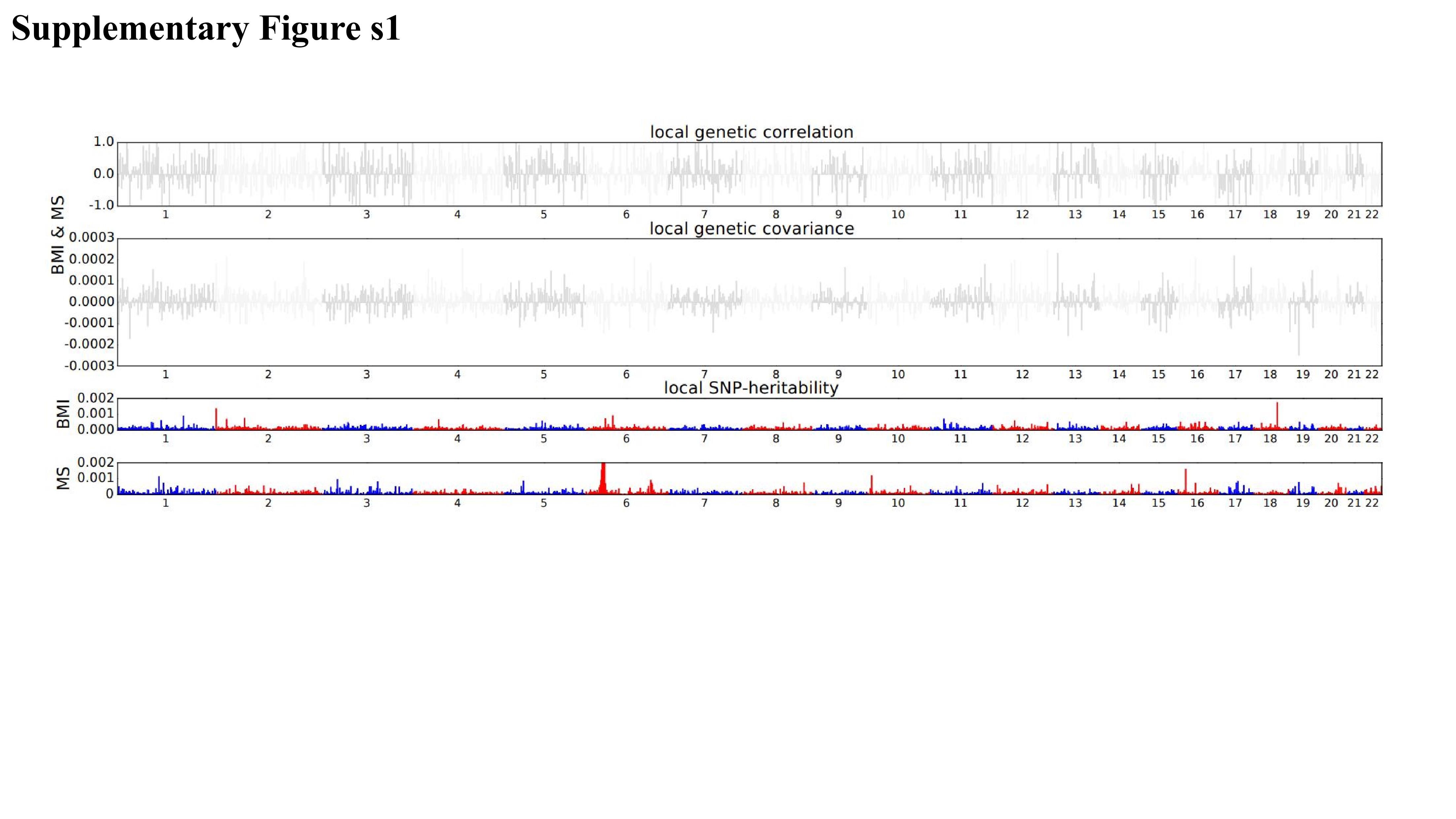

### Supplementary Figure S2

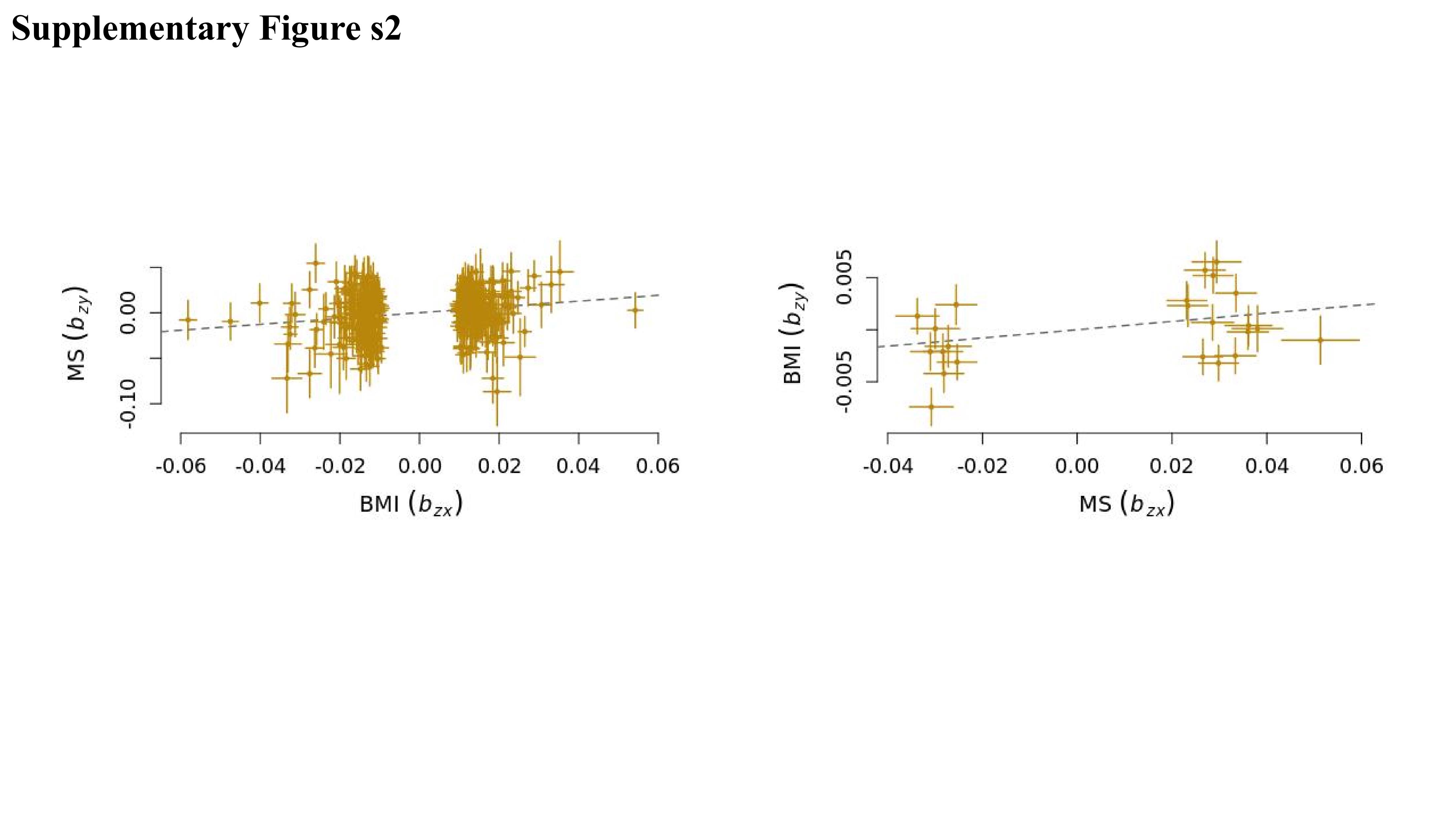

### Supplementary Figure S3

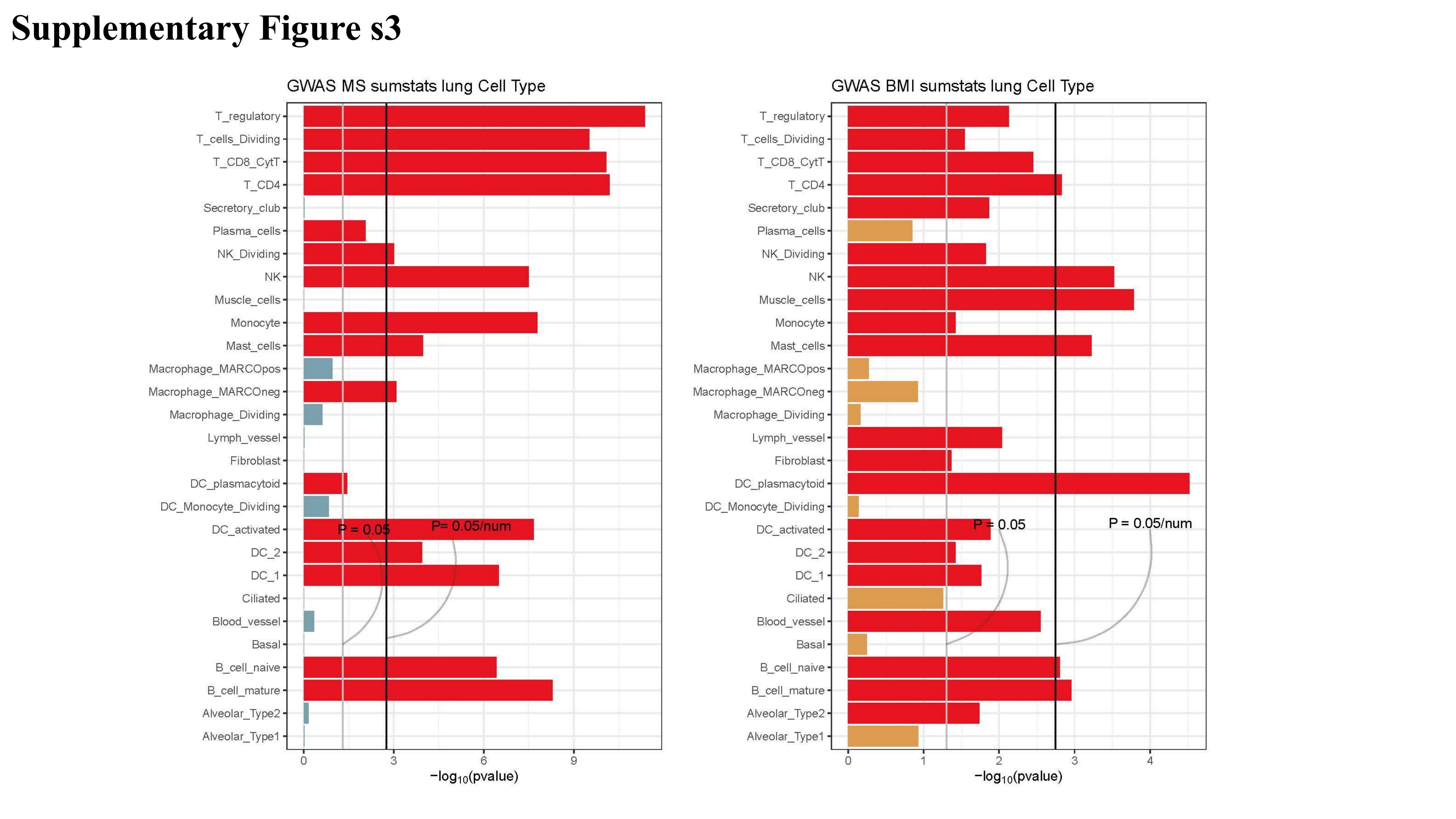

### Supplementary Figure S4

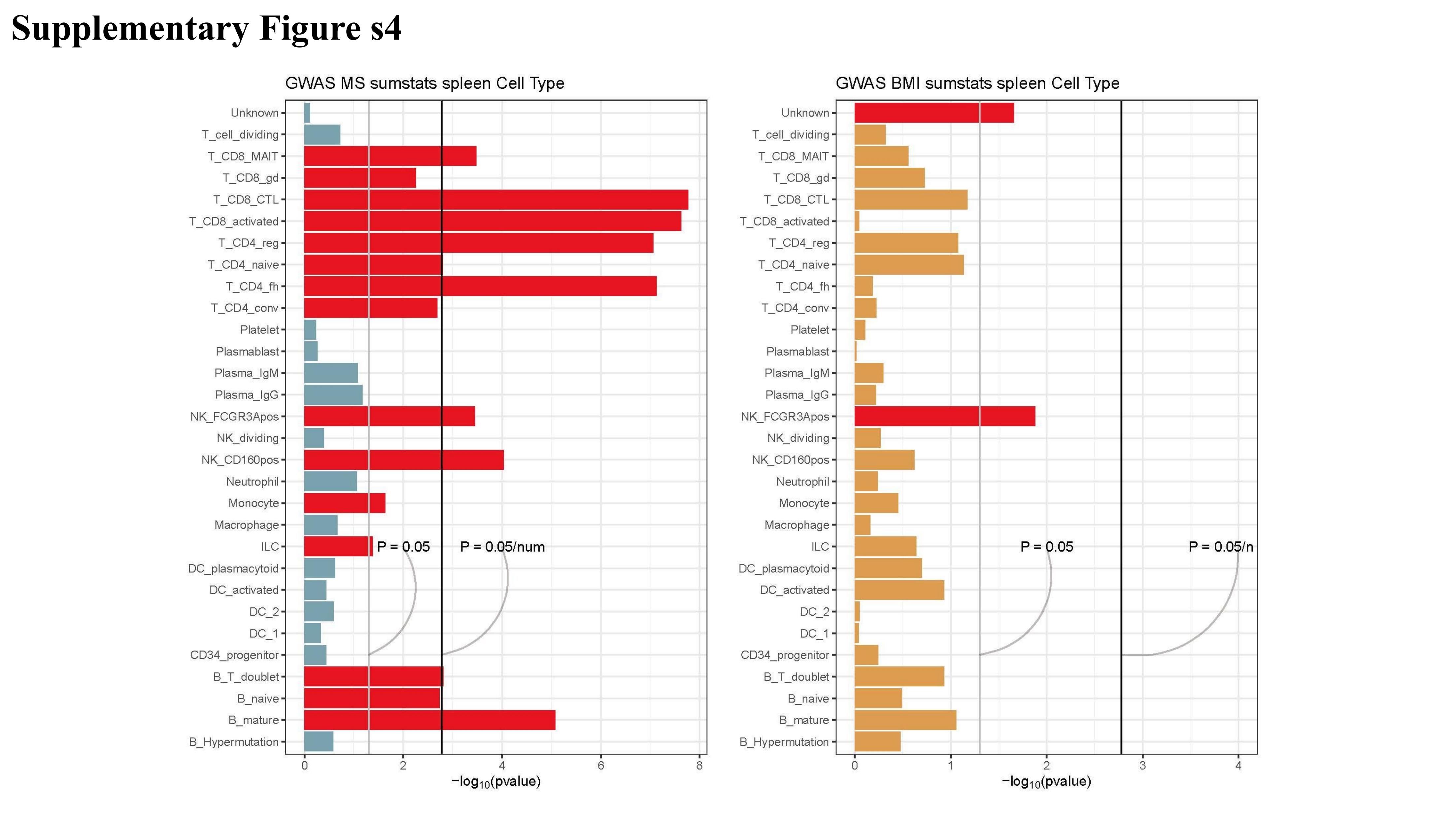

### Supplementary Figure S5

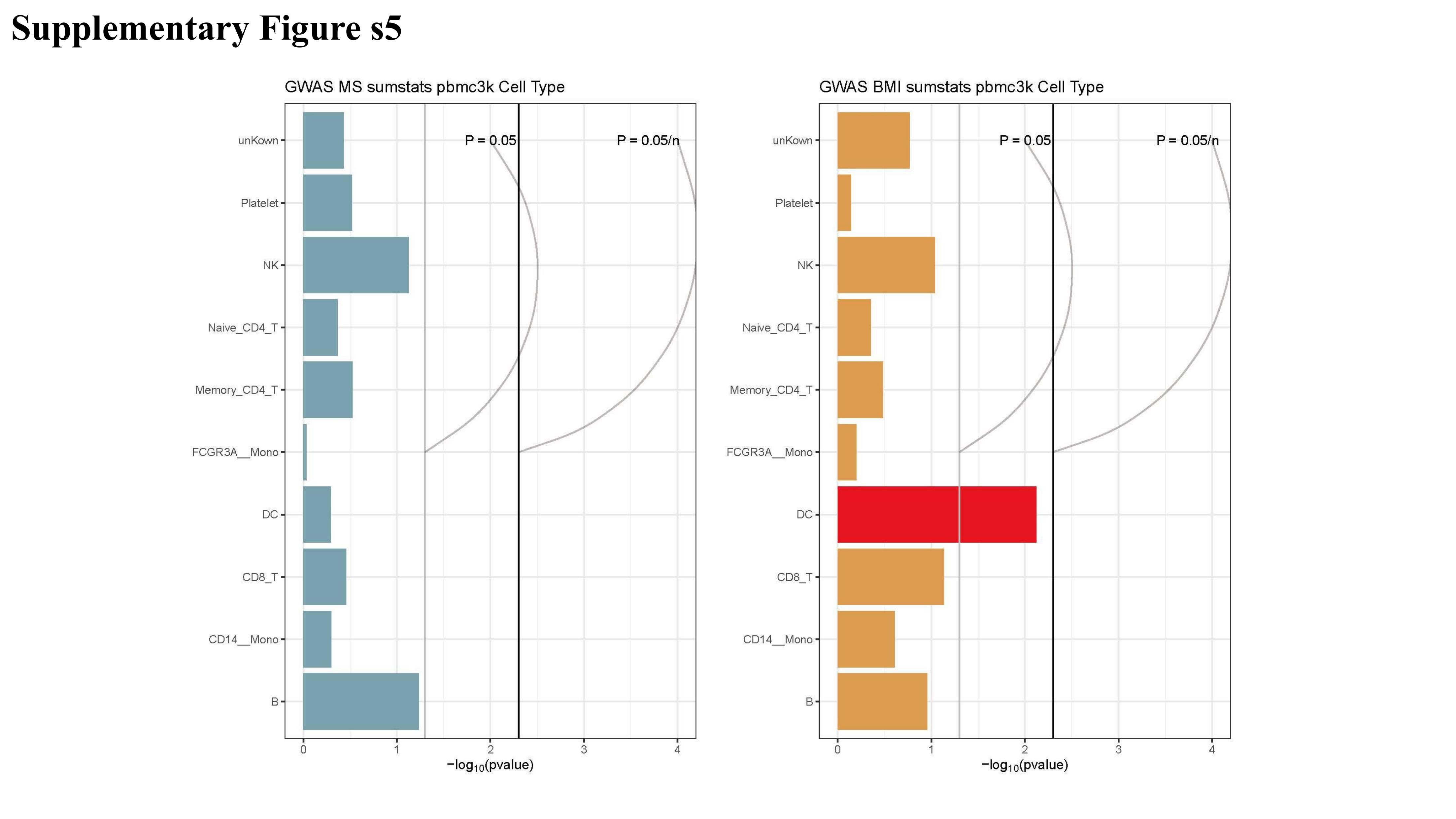

### Supplementary Figure S6

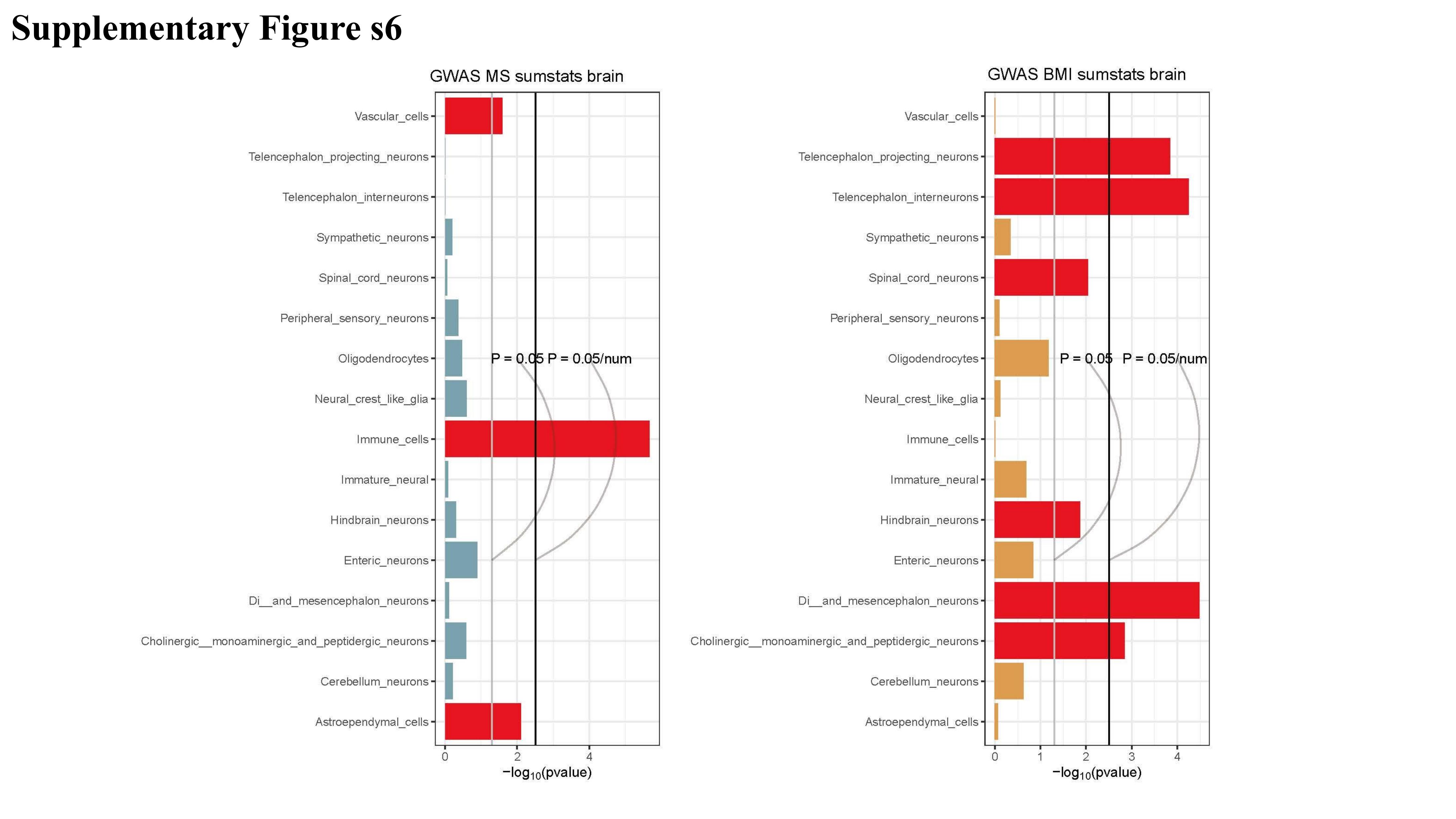
