## Supplementary Table S1-S3 for "Investigating shared genetic architecture between obesity and multiple sclerosis"

**Supplement Table**

**Supplemental Table S1**. **Heritability of BMI and MS and their genetic correlation estimated using LDSC, GNOVA and ρ-HESS.**

|  | **Method** | **Body mass index** | **Multiple sclerosis** |
| --- | --- | --- | --- |
| heritability h^2^ | LDSC | 0.2119 | 0.0457 |
| heritability h^2^ | GNOVA | 0.1885 | 0.1385 |
| heritability h^2^ | ρ-HESS | 0.224 | 0.235 |
| Genetic correlation *r*_g_ | LDSC | 0.0796 | |
| Genetic correlation *r*_g_ | GNOVA | 0.0647 | |
| Genetic correlation *r*_g_ | ρ-HESS | 0.0425 | |

Heritability of BMI and MS and their genetic correlation estimated using LDSC, GNOVA and ρ-HESS. LD: linkage disequilibrium. GNOVA: Genetic covariance analyzer; ρ-HESS: Heritability Estimation from Summary Statistics.

**Supplement Table S2** **Summary of five MR results between BMI and MS**

| BMI to MS | | | | |
| --- | --- | --- | --- | --- |
| Method | SNPs | BETA | SE | *P*-value |
| MR Egger | 436 | 0.203553 | 0.160343 | 0.205 |
| Weighted median | 436 | 0.2026 | 0.090419 | 0.025 |
| Inverse variance weighted | 436 | 0.229651 | 0.056436 | 4.72E-05 |
| Weighted mode | 436 | 0.222631 | 0.180782 | 0.219 |
| GSMR | 380(368) | 0.3194061 | 0.049 | 6.15E-13 |
| MS to BMI | | | | |
| Method | SNPs | BETA | SE | *P*-value |
| MR Egger | 15 | -0.02671 | 0.024555 | 0.296397 |
| Weighted median | 15 | 1.46E-06 | 0.005744 | 0.999796 |
| Inverse variance weighted | 15 | 0.002541 | 0.004214 | 0.54648 |
| Weighted mode | 15 | -0.00028 | 0.008872 | 0.975564 |
| GSMR | 24(23) | 0.03947219 | 0.01229873 | 0.001329881 |

Numbers in the brackets of GSMR results are the numbers of SNPs remaining after the HEIDI (Heterogeneity In Dependent Instrument) test. SNP: single nucleotide polymorphisms. BETA: the estimated effect of exposure on outcome. SE: standard error;

**Supplemental Table S3**.  **Genetic correlations between BMI and MS. Trait1 and Trait2 represent BMI and MS respectively.**

| **With constrained intercept of heritability** | | | | | | |
| --- | --- | --- | --- | --- | --- | --- |
| **r_g_(se)** | *P*-value | Genetic covariance (se) | Traut1 Lambda GC | Traut1 Intercept | Traut2 | Traut2 |
|  |  |  |  |  | Lambda GC | Intercept |
| **0.0806 (0.0215)** | 0.0002 | 0.0117 (0.0031) | 2.7872 | 1.0191 (0.0258) | 1.1491 | 1.0221 (0.0108) |
| **Without constrained intercept** | | | | | | |
| **r_g_(se)** | *P*-value | Genetic covariance (se) | Traut1 Lambda GC | Traut1 Intercept | Traut2 | Traut2 |
|  |  |  |  |  | Lambda GC | Intercept |
| **0.0786 (0.0142)** | 3.45×10-8 | 0.0121 (0.0021) | 2.7872 | 1 | 1.1491 | 1 |
